# Right Ventricular Dysfunction in Ventilated Patients with COVID-19 (COVID-RV)

**DOI:** 10.1101/2021.07.29.21261190

**Authors:** Philip McCall, Jennifer Willder, Bethany Stanley, Claudia-Martina Messow, John Allan, Lisa Gemmell, Alex Puxty, Dominic Strachan, Colin Berry, Ben Shelley

## Abstract

**Purpose:** COVID-19 is associated with cardiovascular complications, with right ventricular dysfunction (RVD) commonly reported. The combination of acute respiratory distress syndrome (ARDS), injurious invasive ventilation, micro/macro thrombi and the potential for direct myocardial injury create conditions where RVD is likely to occur. No study has prospectively explored the prevalence of RVD, and its association with mortality, in a cohort requiring mechanical ventilation.

**Methods:** Prospective, multi-centre, trans-thoracic echocardiographic, cohort study of ventilated patients with COVID-19 in Scottish intensive care units. RVD was defined as the presence of severe RV dilatation and interventricular septal flattening. To explore role of myocardial injury, high sensitivity troponin and N-terminal pro B-type natriuretic peptide (NT-proBNP) were measured in all patients.

**Results:** One hundred and twenty-one patients were recruited to COVID-RV, 118 underwent imaging and it was possible to determine the primary outcome in 112. RVD was present in seven (6.2% [95%CI; 2.5%, 12.5%]) patients. Thirty-day mortality was 85.7% in those with RVD, compared to 44.8% in those without (p=0.051). Patients with RVD were more likely to have; pulmonary thromboembolism (p<0.001), higher plateau pressure (p=0.048), lower dynamic compliance (p=0.031), higher NT-proBNP (p<0.006) and more frequent *abnormal* troponin (p=0.048). Abnormal NT-proBNP (OR 4.77 [1.22, 21.32], p=0.03) and abnormal Troponin (16.54 [4.98, 67.12], p<0.001) independently predicted 30-day mortality.

**Conclusion:** COVID-RV demonstrates a prevalence of RVD in ventilated patients with COVID-19 of 6.2% and is associated with a mortality of 85.7%. Association is observed between RVD and each of the aetiological domains of; ARDS, ventilation, micro/macro thrombi and myocardial injury.

## INTRODUCTION

Since March 2020 there has been a global pandemic of Coronavirus disease 2019 (COVID-19), caused by the Severe Acute Respiratory Syndrome Coronavirus-2 (SARS-CoV-2). COVID-19 can result in acute hypoxaemic respiratory failure which in severe cases may require admission to intensive care (ICU) and invasive mechanical ventilation. Haemodynamic instability and cardiac complications are prevalent, with right ventricular dysfunction (RVD) a common finding [1–10]. The combination of acute respiratory distress syndrome (ARDS), injurious invasive ventilation, micro/macro thrombi and the potential for direct myocardial injury create a *perfect storm* of pathophysiology where RVD is likely to occur [11]. In reports pre-dating the COVID-19 pandemic, RVD occurs in up to 55% of patients with ARDS and is independently associated with mortality, with increasing ARDS severity associated with increased frequency of RV dysfunction [12,13].

Previous echocardiography studies in COVID-19 have demonstrated RVD and association with adverse clinical outcomes [1,3–10]. To date however, most of the research into RVD in COVID-19 has been retrospective and performed in undifferentiated cohorts, *of ventilated and non-ventilated patients* [3–6,8–10]. As highlighted by Michard and Vieillard-Baron in February’s issue of *Intensive Care Medicine*, to date, there have been no robust, *prospective*, haemodynamic evaluations of patients with COVID-19 focusing *specifically on the intensive care unit population* [2]. To this end, COVID-RV was designed to prospectively investigate the prevalence of RV dysfunction in ventilated patients with COVID-19, any association with mortality and, to explore causative mechanisms.

## METHODS

### Study design and participants

Study protocol and methods have previously been reported [14]. We conducted a prospective, observational cohort study involving 10 ICUs in NHS Scotland. Ethics approval was obtained from Scotland A Research Ethics Committee (responsible for studies requiring approval under the Adults with Incapacity [Scotland] Act, 2000 - 20/SS/0059). Patients were eligible for inclusion if they were more than 16 years old with confirmed SARS-CoV-2 infection, with severe acute respiratory failure requiring tracheal intubation and positive pressure ventilation in ICU for more than 48 hours, but not more than 14 days. Exclusion criteria were; pregnancy, ongoing participation in investigational research that may undermine the scientific basis of the study, prior participation in COVID-RV, requirement for extracorporeal membrane oxygenation support (for respiratory or cardiovascular failure) and end of life care where the patient was not expected to survive longer than 24 hours. COVID-RV was registered at ClinicalTrials.gov (NCT04764032).

### Data

Study data were collected and managed using REDCap electronic data capture tools hosted by the University of Glasgow.

#### Clinical and laboratory data

Baseline demographics, chronic comorbidities, clinical trajectory prior to ICU admission, severity of illness, acute comorbidities and follow-up data were all collected prospectively. Clinical data relating to potential mechanisms of RV dysfunction were also collected, specifically regarding the four domains of; ARDS, disordered coagulation, myocardial injury and mechanical ventilation.

#### Echocardiography

Participants underwent a single transthoracic echocardiogram (TTE) to determine the presence or absence of RV dysfunction. To reflect the clinical practice of bedside echocardiography in intensive care, for the purposes of determining the primary outcome of the study, imaging required was in keeping with the protocol required for a focused intensive care echo (FICE) scan [15]. In line with previous reports, RVD was defined as TTE evidence of severe RV dilatation along with the presence of interventricular septal flattening. Severe RV dilatation was determined from the apical 4-chamber view at end diastole and was present when the RV:left ventricular (LV) ratio was >1.

#### Cardiac biomarkers

High sensitivity troponin (I or T depending on assay used at each site) and N-terminal pro B-type natriuretic peptide (NT-proBNP) were measured in all patients on the day of echocardiography. Samples were processed alongside routine clinical samples in each host site, and therefore subject to routine laboratory quality assurance processes. Abnormal values were defined for NT-proBNP (>300ng/ml) and Troponin (TnT ≥15 ng/L or TnI ≥34 ng/L for males; ≥16 ng/L for females).

### Statistical considerations

Statistical analyses were performed by statisticians (B.S., C.-M.M.) based in the Robertson Centre for Biostatistics at the University of Glasgow.

#### Outcomes

The primary outcome was the prevalence of RV dysfunction and its association with 30-day mortality. Exploratory outcomes sought to determine association between RVD and proposed aetiological factors. Additionally, association between cardiac biomarker levels and 30-day mortality was assessed.

#### Power

Sample size selection was, by necessity, a pragmatic balance of maximising available information versus the prompt delivery of the study. Given the number of patients admitted to ICU in Scotland during the first wave of the pandemic (Q2, 2020), we believed it was realistic to recruit 120–150 patients across participating sites. Power calculations were performed for estimated RV dysfunction prevalence rates of 25% and 50%, with an overall mortality rate of 50% and are demonstrated in supplementary table 1. These analyses suggest that a study of 120 patients (the ultimate sample size) would have 80% power to detect an associated odds ratio (OR) for mortality of 2.83 to 3.43 (with an estimated prevalence of RVD of 50% and 25% respectively).

#### Statistical methods

The proportion of ventilated patients with COVID-19 who have RVD was determined, with a 95% confidence interval (CI) utilising the Clopper-Pearson method. We then sought to analyse the association of RV dysfunction with 30-day mortality using logistic regression analysis predicting 30-day mortality from presence or absence of RV dysfunction, adjusting for patient demographics (age, gender, ethnicity), phase of disease (time from intubation to echocardiography) and baseline severity of illness (Acute Physiology And Chronic Health Evaluation II [APACHE II] score). Firth’s bias-reduced logistic regression was used where ordinary logistic regression failed due to small numbers [16]. Association of the cardiac biomarkers with 30-day mortality was assessed using logistic regression analysis, predicting 30-day mortality and adjusted for patient demographics, phase of disease and baseline severity of illness. Ordinal and categorical data are presented as n (%). Continuous data are summarised as mean (standard deviation [SD]) or median (interquartile range [IQR]) as appropriate. Between-group differences were assessed using Fisher’s Exact test for categorical variables and Student’s T- or Wilcoxon Mann-Whitney test for continuous variables. Statistical analyses were performed using R version 4.0.0 (R Foundation for Statistical Computing, Vienna, Austria).

## RESULTS

Between 2^nd^ September 2020 and 22^nd^ March 2021, 121 patients were recruited to COVID-RV (Figure 1). Following recruitment, three patients were excluded from further participation; one who was extubated prior to echocardiography and two as a result of technical factors preventing imaging. Patient characteristics at ICU admission are provided in table 1. Thirty-day mortality in the whole cohort was 47.3% (53 of 112 died).

**Table 1.** Patient characteristics at ICU admission.

| Table 1. Patient characteristics at ICU admission |  |  |  |  |  |
| --- | --- | --- | --- | --- | --- |
|  |  | All<br>(n=112) | RVD<br>(n=7) | No RVD<br>(n=105) | p-value |
| Age, years |  | 59.2 (11.3) | 61.1 (4.4) | 59 (11.6) | 0.318<sup>#</sup> |
| Male |  | 74 (66.1%) | 5 (71.4%) | 69 (65.7%) | >0.99<sup>†</sup> |
| BMI, kg/m2 | *n (n missing)* | 110 (2) | 7 (0) | 103 (2) |  |
|  |  | 32.9 (7.1) | 31.5 (4.6) | 32.9 (7.2) | 0.467<sup>#</sup> |
| Ethnicity | White | 100 (89.3%) | 7 (100%) | 93 (88.6%) |  |
|  | Non-white | 12 (10.7%) | 0 (0%) | 1 (1%) | >0.99<sup>†</sup> |
| Time since symptom onset to intubation, days |  | 11 [7, 16] | 18 [13, 25] | 10.5 [7, 14.2] | 0.035<sup>‡</sup> |
| Clinical frailty score | *n (n missing)* | 111 (1) | 7 (0) | 104 (1) |  |
|  |  | 2.4 (0.9) | 2 (0.8) | 2.4 (0.9) | 0.281<sup>#</sup> |
| APACHE II | *n (n missing)* | 107 (5) | 7 (0) | 100 (5) |  |
|  |  | 16.6 (5.8) | 17.6 (6.1) | 16.6 (5.8) | 0.687<sup>#</sup> |
| CCCC | *n (n missing)* | 102 (10) | 7 (0) | 95 (10) |  |
|  |  | 10.2 (2.7) | 10.6 (2.3) | 10.2 (2.8) | 0.712<sup>#</sup> |
| Comorbidities |  |  |  |  |  |
| Smoking | Non-smoker | 63 (56.2%) | 2 (28.6%) | 61 (58.1%) |  |
|  | Ex-smoker > 1 year | 40 (35.7%) | 4 (57.1%) | 36 (34.3%) | =0.228<sup>†</sup> |
|  | Current or within 1 year | 9 (8%) | 1 (14.3%) | 8 (7.6%) |  |
| Alcohol history | *n (n missing)* | 110 (2) | 7 (0) | 103 (2) |  |
|  | None | 34 (30.9%) | 1 (14.3%) | 33 (32%) |  |
|  | Minimal | 57 (51.8%) | 6 (85.7%) | 51 (49.5%) | =0.478<sup>†</sup> |
|  | Moderate | 8 (7.3%) | 0 (0%) | 8 (7.8%) |  |
|  | Excess | 11 (10%) | 0 (0%) | 11 (10.7%) |  |
| Hypertension |  | 38 (33.9%) | 2 (28.6%) | 36 (34.3%) | >0.99<sup>†</sup> |
| Coronary artery disease |  | 11 (9.8%) | 0 (0%) | 11 (10.5%) | >0.99<sup>†</sup> |
| Diabetes |  | 33 (29.5%) | 2 (28.6%) | 31 (29.5%) | >0.99<sup>†</sup> |
| Asthma |  | 16 (14.3%) | 1 (14.3%) | 15 (14.3%) | >0.99<sup>†</sup> |
| COPD |  | 10 (8.9%) | 0 (0%) | 10 (9.5%) | >0.99<sup>†</sup> |
| Treatments before intubation |  |  |  |  |  |
| Intravenous corticosteroids |  | 74 (66.1%) | 4 (57.1%) | 70 (66.7%) | 0.687<sup>†</sup> |
| Non-invasive ventilation |  | 76 (67.9%) | 6 (85.7%) | 70 (66.7%) | 0.426<sup>†</sup> |
| High flow nasal oxygen |  | 65 (58%) | 6 (85.7%) | 59 (56.2%) | 0.235<sup>†</sup> |
| Awake self-proning |  | 57 (50.9%) | 6 (85.7%) | 51 (48.6%) | 0.114<sup>†</sup> |
| Acute comorbidities since hospital admission |  |  |  |  |  |
| New arrhythmias |  | 17 (15.2%) | 3 (42.9%) | 14 (13.3%) | 0.070<sup>†</sup> |
| Confirmed or suspected PTE | Radiologically confirmed | 4 (3.6%) | 3 (42.9%) | 1 (1%) |  |
|  | Clinically suspected | 5 (4.5%) | 1 (14.3%) | 4 (3.8%) |  |
|  | No | 101 (90.2%) | 3 (42.9%) | 98 (93.3%) | <0.001<sup>†</sup> |
|  | Unknown | 2 (1.8%) | 0 (0%) | 2 (1.9%) |  |
| ACS |  | 5 (4.5%) | 0 (0%) | 5 (4.8%) | >0.99<sup>†</sup> |
| Requirement for RRT |  | 18 (16.1%) | 2 (28.6%) | 16 (15.2%) | 0.313<sup>†</sup> |

Data are presented as mean (SD), median [IQR] or n (%). Data are complete unless indicated by *n (n missing)*. P-values provided are for comparisons between RVD vs no RVD. # = Student's T-test. † = Fisher's Exact test. ‡ = Wilcoxon Mann-Whitney test.
RVD = Right Ventricular Dysfunction, BMI = Body Mass Index, APACHE = Acute Physiology And Chronic Health Evaluation, CCCC = Coronavirus Clinical Characterisation Consortium, COPD = Chronic Obstructive Pulmonary Disease, PTE = Pulmonary Thromboembolism, ACS = Acute Coronary Syndrome, RRT = Renal Replacement Therapy.

**Figure 1.**
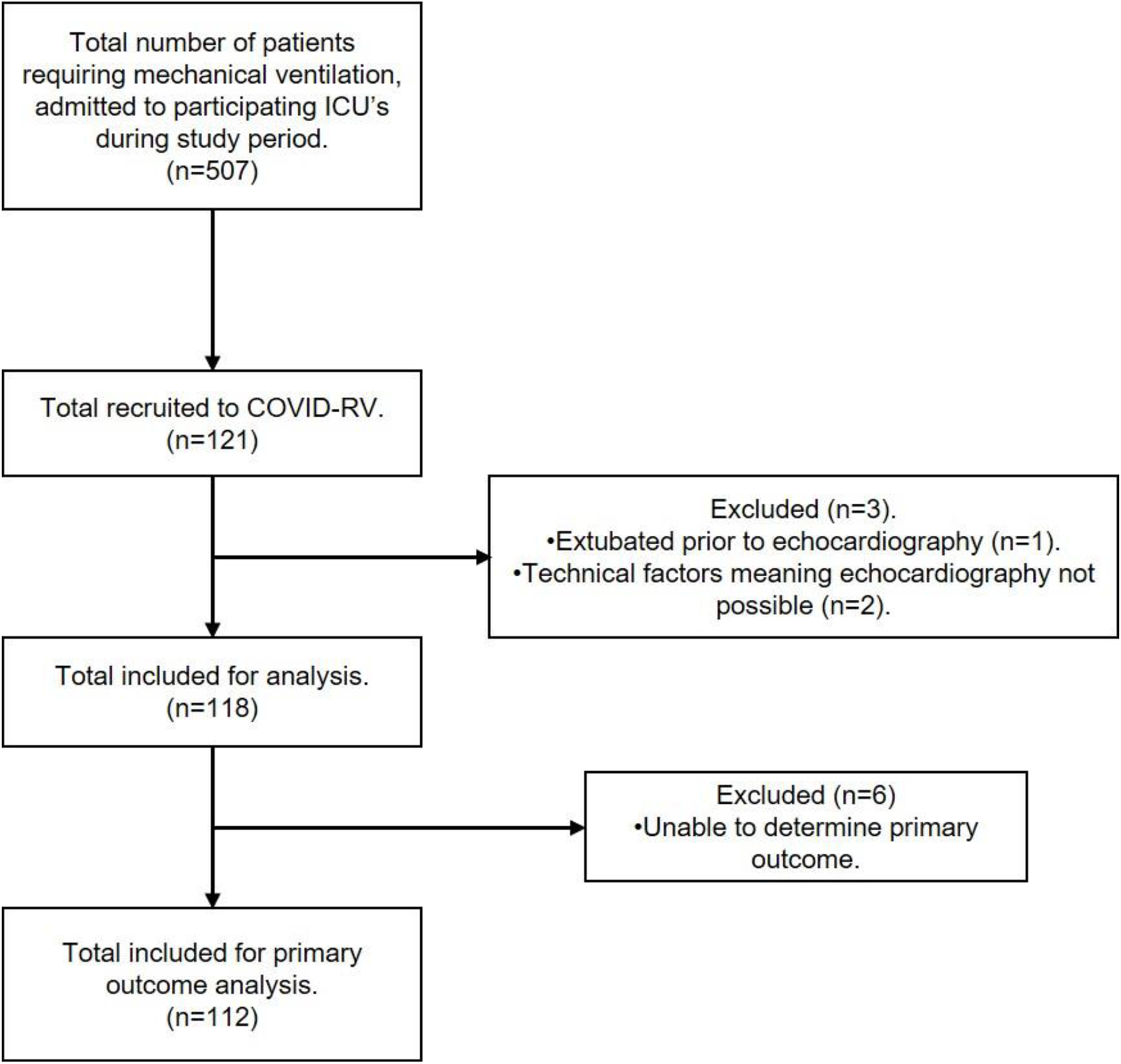
Patient recruitment.

Of the 118 patients where TTE was performed, it was possible to determine the primary outcome in 112 (94.9%, supplementary table 1). Echocardiography was performed by; British Society of Echocardiography accredited echocardiographers (n=55 [46.6%]), FICE accredited critical care clinicians with “mentor” status (n=37 [31.4%]), FICE accredited clinicians (n=8 [6.8%]) and clinicians without any formal accreditation (n=18 [15.3%]). Imaging was performed a median of 5 (4, 8) days following intubation. Thirty-one (27.7%) patients had evidence of severe RV dilatation (RV:LV ratio >1:1 on A4C view) and nine (8%) had evidence of interventricular septal flattening. Right ventricular dysfunction (the combination of these two parameters and primary endpoint) was present in seven (6.2% [95% CI; 2.5%, 12.5%]) patients (Figure 2). *Subjective* RV dysfunction was present in 85.7% of those with RVD, in contrast to 9.6% in those without (p<0.001). *Subjective* LV dysfunction was present in 28.6% of those with RVD, in contrast to 9.7% in those without (p=0.168, table 2). There was no difference in time from *intubation* to imaging in those with or without RVD (p=0.942); time from *symptom onset* to echocardiography however was longer in those with RVD compared to those without (23 [20, 29.5] days compared to 17 [13, 21.2] days, p=0.017, table 2).

**Table 2.** Echocardiography.

|  |  | **All**<br>**(n=112)** | **RVD**<br>**(n=7)** | **No RVD**<br>**(n=105)** | **p-value** |
| --- | --- | --- | --- | --- | --- |
| **Time from symptom onset to echocardiography, days** | *n (n missing)* | 111 (1)<br>18 [13, 22] | 7 (0)<br>23 [20, 29.5] | 104 (1)<br>17 [13, 21.2] | 0.017<sup>‡</sup> |
| **Time from intubation to echocardiography, days** |  | 5 [4, 8] | 5 [5, 6] | 5 [4, 8] | 0.942<sup>‡</sup> |
| **RV dilatation** | *n (n missing)* | 110 (2)<br>31 (27.7%) | 7 (0)<br>7 (100%) | 103 (2)<br>24 (22.9%) | - |
| **Septal flattening** | *n (n missing)* | 109 (3)<br>9 (8%) | 7 (0)<br>7 (100%) | 102 (3)<br>2 (1.9%) | - |
| **Subjective LV dysfunction** | *n (n missing)* | 110 (2)<br>12 (10.9%) | 7 (0)<br>2 (28.6%) | 103 (2)<br>10 (9.7%) | 0.168<sup>†</sup> |
| **Subjective RV dysfunction** | *n (n missing)* | 111 (1)<br>16 (14.4%) | 7 (0)<br>6 (85.7%) | 104 (1)<br>10 (9.6%) | <0.001<sup>†</sup> |

Data are presented as median [IQR] or n (%). Data are complete unless indicated by *n (n missing)*. P-values provided are for comparisons between RVD vs no RVD. <sup>‡</sup> = Wilcoxon Mann-Whitney test. <sup>†</sup> = Fisher's Exact test. RV = Right Ventricular, LV = Left Ventricular.

**Figure 2.**
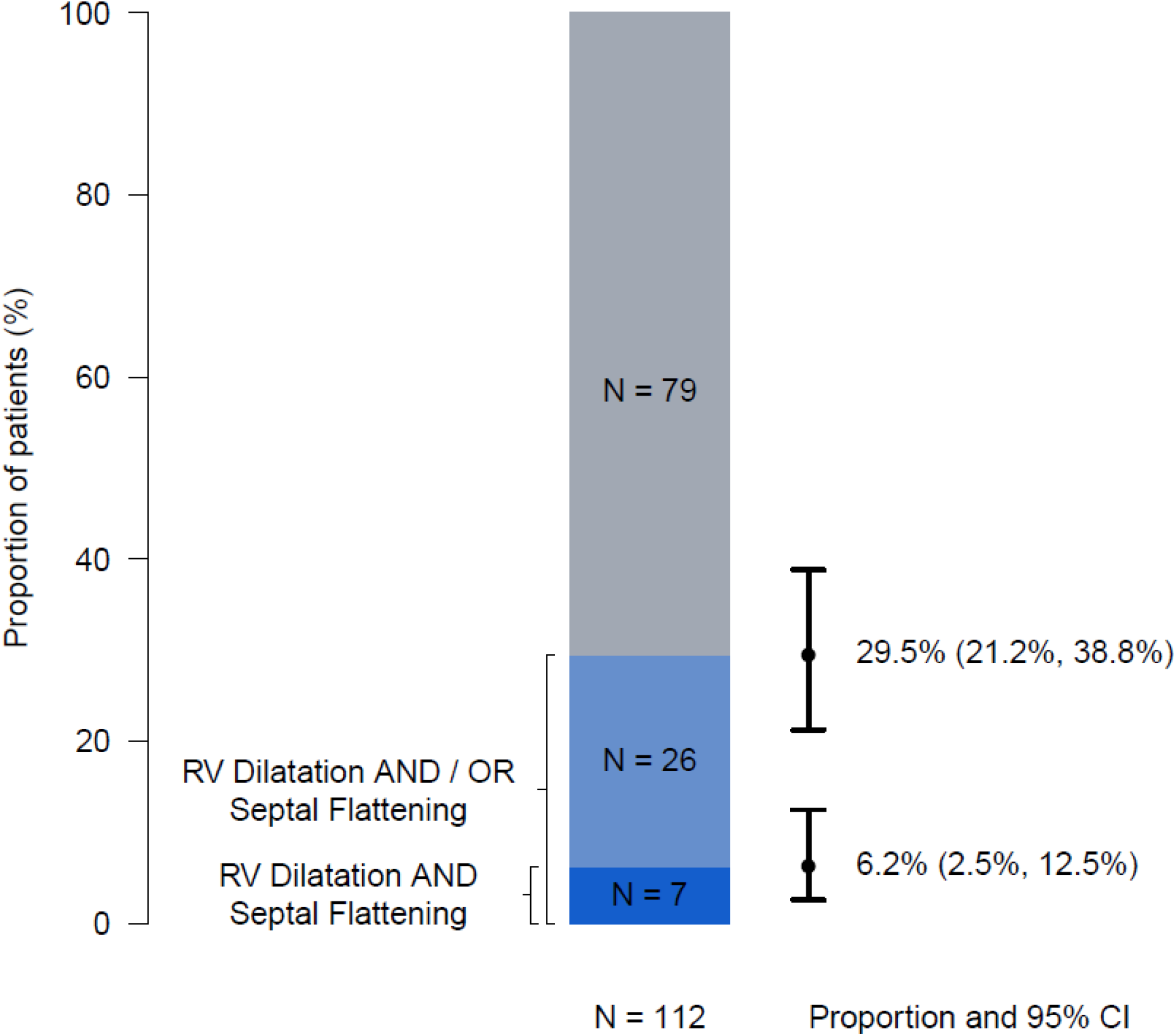
Proportion of patients with Right Ventricular Dysfunction.

Patients with RVD had a higher creatinine at the time of imaging and were more likely to be undergoing RRT (p=0.014 and 0.049 respectively, table 5). Heart rate was higher in patients with RVD (p=0.016) and they had more disordered acid-base status (p≤0.001, table 5).

**Table 3.** Clinical consequences.

|  | **All**<br>**(n=112)** | **RVD**<br>**(n=7)** | **No RVD**<br>**(n=105)** | **p-value** |
| --- | --- | --- | --- | --- |
| *30-day follow-up* |  |  |  |  |
| **Death** | 53 (47.3%) | 6 (85.7%) | 47 (44.8%) | 0.051<sup>†</sup> |
| **RRT** | 28 (25%) | 3 (42.9%) | 25 (23.8%) | 0.581<sup>†</sup> |
| **Prone ventilation** | 55 (49.1%) | 2 (28.6%) | 53 (50.5%) | 0.322<sup>†</sup> |
| **Referral for ECMO** | 15 (13.4%) | 0 (0.0%) | 15 (14.3%) | 0.663<sup>†</sup> |

Data are presented as n (%). Data are complete unless indicated by *n* (*n missing*). P-values provided are for comparisons between RVD vs no RVD. † = Fisher's Exact test.
RRT = Renal Replacement Therapy, ECMO = Extra Corporeal Membrane Oxygenation.

**Table 4.** Firth’s bias reduced logistic regression predicting 30-day mortality adjusting for remaining variables in table.

|  | **OR (95% CI)** | **p-value** |
| --- | --- | --- |
| **RVD (RV dilatation *and* septal flattening)** | 5.12 (0.99, 51.38) | =0.051 |
| **Age, in years (per 5-year increase)** | 1.48 (1.19, 1.91) | <0.001 |
| **Female Gender** | 1.12 (0.46, 2.73) | =0.803 |
| **Non-white ethnicity** | 0.92 (0.22, 3.54) | =0.899 |
| **APACHE II score on admission to ICU (per 5-score increase)** | 1.27 (0.87, 1.90) | =0.221 |
| **Time from intubation to date of echo, in days (per 1-day increase)** | 1.01 (0.88, 1.17) | =0.837 |
| RVD = Right Ventricular Dysfunction, APACHE = Acute Physiology And Chronic Health Evaluation |  |  |

**Table 5.** Patient characteristics on day of echocardiography.

|  |  | All<br>(n=112) | RVD<br>(n=7) | No RVD<br>(n=105) | p-value |
| --- | --- | --- | --- | --- | --- |
| Requirement for prone invasive ventilation since ICU admission |  | 79 (70.5%) | 7 (100%) | 72 (68.6%) | 0.240<sup>†</sup> |
| SOFA score |  | 7.9 (3) | 9.3 (4.2) | 7.8 (2.9) | 0.408<sup>#</sup> |
| Requirement for RRT on day of echo |  | 15 (13.4%) | 3 (42.9%) | 12 (11.4%) | 0.049<sup>†</sup> |
| *Lab Measurements* |  |  |  |  |  |
| *Arterial Blood Gas* |  |  |  |  |  |
| [H<sup>+</sup>], nmol/L) | *n (n missing)* | 97 (15)<br>39 [35.8, 46] | 5 (2)<br>53 [53, 60.9] | 92 (13)<br>38.5 [35, 45] | =0.001<sup>‡</sup> |
| PaO<sub>2</sub>, kPa |  | 9.3 (1.2) | 8.8 (1) | 9.3 (1.3) | 0.219<sup>#</sup> |
| PaCO<sub>2</sub>, kPa | *n (n missing)* | 109 (3)<br>6.9 [5.9, 8] | 7 (0)<br>7.6 [6.2, 8.8] | 102 (3)<br>6.8 [5.9, 7.9] | 0.390<sup>‡</sup> |
| BE, mmol | *n (n missing)* | 110 (2)<br>5.9 (6.5) | 7 (0)<br>-1.2 (3.2) | 103 (2)<br>6.4 (6.4) | <0.001<sup>#</sup> |
| Bicarbonate, mmol/L | *n (n missing)* | 109 (3)<br>31.8 (6.6) | 7 (0)<br>26.2 (2.7) | 102 (3)<br>32.1 (6.6) | <0.001<sup>#</sup> |
| *Full Blood Count* |  |  |  |  |  |
| Haemoglobin, g/dl |  | 11 (1.8) | 10.1 (1.2) | 11.1 (1.8) | 0.073<sup>#</sup> |
| Neutrophils, x10<sup>9</sup>/L |  | 10.5 [8.5, 14.9] | 9.8 [8.4, 13.3] | 10.5 [8.5, 14.6] | 0.512<sup>‡</sup> |
| Lymphocytes, x10<sup>9</sup>/L |  | 0.9 [0.5, 1.4] | 1 [0.9, 1.9] | 0.9 [0.5, 1.3] | 0.341<sup>‡</sup> |
| Platelets, x10<sup>9</sup>/L |  | 279.5 (109.5) | 243 (148.2) | 282 (106.9) | 0.518<sup>#</sup> |
| *Inflammation* |  |  |  |  |  |
| CRP, mg/L |  | 61.5 [11.8, 157.5] | 71 [29, 185.5] | 60 [10, 156] | 0.528<sup>‡</sup> |
| *Coagulation* |  |  |  |  |  |
| D-Dimers, mg/L FEU | *n (n missing)* | 81 (31)<br>1264 [601, 2605] | 5 (2)<br>1156 [1105, 1485] | 76 (29)<br>1315 [573, 2651.5] | 0.961<sup>‡</sup> |
| PT, seconds |  | 12 [11, 13.2] | 13 [11.4, 14.5] | 12 [11, 13.2] | 0.434<sup>‡</sup> |
| APTT, seconds | *n (n missing)* | 111 (1)<br>27 [25, 31] | 7 (0)<br>31 [27.1, 35.5] | 104 (1)<br>26.9 [25, 30] | 0.120<sup>‡</sup> |
| *Electrolytes* |  |  |  |  |  |
| Creatinine, µmol/L |  | 69.5 [53.5, 107] | 157 [112, 211.5] | 67 [52, 104] | 0.014<sup>‡</sup> |
| *Cardiac biomarkers* |  |  |  |  |  |
| NT-proBNP, ng/L | *n (n missing)* | 100 (12)<br>458 [198.5, 1689.5] | 7 (0)<br>4806 [2571.5, 17121] | 93 (12)<br>429 [197, 1369] | 0.006<sup>‡</sup> |
| Abnormal NT-proBNP<sup>A</sup> | *n (n missing)* | 100 (12)<br>63 (63%) | 7 (0)<br>6 (85.7%) | 93 (12)<br>57 (61.3%) | 0.255<sup>†</sup> |
| hsTn I, ng/L | *n (n missing)* | 64 (48)<br>12 [4.8, 42] | 5 (2)<br>56 [38, 102] | 59 (46)<br>11 [4, 37] | 0.082<sup>‡</sup> |
| hsTn T, ng/L | *n (n missing)* | 46 (66)<br>16.5 [10, 28.5] | 2 (5)<br>39 [34, 44] | 44 (61)<br>16 [10, 26.2] | 0.094<sup>‡</sup> |
| Abnormal troponin<sup>B</sup> | *n (n missing)* | 110 (2)<br>51 (46.4%) | 7 (0)<br>6 (85.7%) | 103 (2)<br>45 (43.7%) | 0.048<sup>†</sup> |
| *Clinical Parameters* |  |  |  |  |  |
| HR, bpm |  | 79 (19.9) | 99.1 (17.3) | 77.7 (19.5) | 0.016<sup>#</sup> |
| Rhythm | Sinus<br>AF/Flutter | 107 (95.5%)<br>5 (4.5%) | 7 (100%)<br>0 (0%) | 100 (95.2%)<br>5 (4.8%) | >0.99<sup>†</sup> |
| Mean BP, mmHg | *n (n missing)* | 109 (3)<br>80.9 (13.4) | 7 (0)<br>74.1 (9.6) | 102 (3)<br>81.3 (13.5) | 0.100<sup>#</sup> |
| CVP, cmH<sub>2</sub>O | *n (n missing)* | 74 (38)<br>7 [4, 12] | 4 (3)<br>13.5 [10.2, 16.8] | 70 (35)<br>7 [4, 11.8] | 0.187<sup>‡</sup> |
| *Drug Administration* |  |  |  |  |  |
| Vasopressors |  | 40 (35.7%) | 4 (57.1%) | 36 (34.3%) | 0.246<sup>†</sup> |
| Inotropes |  | 1 (0.9%) | 0 (0%) | 1 (1%) | >0.99<sup>†</sup> |
| Anticoagulation | Prophylactic<br>Therapeutic<br>None | 93 (83%)<br>17 (15.2%)<br>2 (1.8%) | 3 (42.9%)<br>4 (57.1%)<br>0 (0%) | 90 (85.7%)<br>13 (12.4%)<br>0 (0%) | 0.018<sup>†</sup> |
| Paralysis |  | 52 (46.4%) | 3 (42.9%) | 49 (46.7%) | >0.99<sup>†</sup> |
| *Ventilation* |  |  |  |  |  |
| FiO<sub>2</sub> |  | 0.6 [0.4, 0.7] | 0.6 [0.5, 0.7] | 0.6 [0.4, 0.7] | 0.659<sup>‡</sup> |
| **Requirement for prone ventilation in previous 24 hours** |  | 44 (39.3%) | 4 (57.1%) | 40 (38.1%) | 0.468<sup>†</sup> |
| **Plateau pressure, cmH<sub>2</sub>O** |  | 58 (54)<br>24 [22, 27] | 5 (2)<br>27 [27, 29] | 53 (52)<br>24 [22, 27] | 0.048<sup>‡</sup> |
| **PAP, cmH<sub>2</sub>O** | *n (n missing)* | 58 (54)<br>25 [20, 30] | 5 (2)<br>29 [27, 30.5] | 53 (52)<br>25 [20, 29] | 0.171<sup>‡</sup> |
| **Tidal volume, ml/kg (PBW)** | *n (n missing)* | 108 (4)<br>7.2 (2.1) | 7 (0)<br>7.1 (3.3) | 101 (4)<br>7.2 (2) | 0.978<sup>#</sup> |
| **P/F ratio** |  | 17 [13.6, 21.3] | 15.6 [12.5, 17.5] | 17.5 [13.7, 21.5] | 0.361<sup>‡</sup> |
| **PEEP, cmH<sub>2</sub>O** | *n (n missing)* | 111 (1)<br>9.8 (3.6) | 7 (0)<br>8.7 (3) | 104 (1)<br>9.9 (3.6) | 0.351<sup>#</sup> |
| **Resp rate (/minute)** |  | 24.3 (5.3) | 25.3 (6.3) | 24.2 (5.2) | 0.676<sup>#</sup> |
| **Driving pressure, cmH<sub>2</sub>O** | *n (n missing)* | 58 (54)<br>14.7 (7.1) | 5 (2)<br>19.4 (5.1) | 53 (52)<br>14.2 (7.1) | 0.086<sup>#</sup> |
| **Dynamic compliance, ml/cmH<sub>2</sub>O** | *n (n missing)* | 58 (54)<br>29.8 [20, 40] | 5 (2)<br>18.8 [17, 20] | 53 (52)<br>32.1 [21.1, 40.6] | 0.031<sup>‡</sup> |
| **Murray lung injury score** | *n (n missing)* | 100 (12)<br>2.8 [2.3, 3] | 6 (1)<br>2.9 [2.6, 3.2] | 94 (11)<br>2.8 [2.3, 3] | 0.491<sup>‡</sup> |

Data are presented as mean (SD), median [IQR] or n (%). Data are complete unless indicated by *n (n missing)*. P-values provided are for comparisons between RVD vs no RVD. # = Student's T-test. † = Fisher's Exact test. ‡ = Wilcoxon Mann-Whitney test. Missing data indicated by *n (n missing)*.
A, NT-proBNP ≥ 300 ng/L
B, hsTnT ≥ 15 ng/L or hsTnI ≥ 34 ng/L for males; ≥ 16 ng/L for females
RVD = Right Ventricular Dysfunction, RRT = Renal Replacement Therapy, BE = Base Excess, CRP = C-Reactive Protein, PT = Prothrombin Time, APTT = Activated Partial Thromboplastin Time, NT-proBNP = N-terminal pro B-type Natriuretic Peptide, hsTn = High Sensitivity Troponin, HR = Heart Rate, BP = Blood Pressure, CVP = Central Venous Pressure, FiO<sub>2</sub> = Fraction of Inspired Oxygen, PAP = Peak Airway Pressure, PBW = Predicted Body Weight, PEEP = Positive End Expiratory Pressure.

### Association with mortality

Six of seven patients (85.7%) with RVD were deceased by the time of 30-day follow up compared to 47 of 105 (44.8%) without RVD (p=0.051, table 3). Firth’s bias-reduced regression analyses demonstrated the presence of RVD was not a significant predictor of 30-day mortality (OR [95%CI] 5.12 [0.99, 51.38], p=0.051, table 4).

### Association with aetiological factors

#### Disordered coagulation

Radiologically confirmed or clinically suspected pulmonary thromboembolism (PTE), was present in 57.1% of patients with RVD, compared to 4.8% in those without (p<0.001, table 1). In keeping with this, treatment with anticoagulation was more common in those with RVD (p=0.018, table 5). There was no difference in platelets, prothrombin time, activated prothrombin time, and D-Dimer between the two groups (p>0.120, table 5).

#### Mechanical ventilation

Where it could be measured, plateau pressure was higher, and compliance lower, in the population with RVD (p=0.048 and 0.031 respectively, table 5). There was no difference in, peak airway pressure, positive end expiratory pressure, driving pressure, or indexed tidal volume between groups (p>0.086 for all, table 5).

#### Severity of ARDS

There was no difference in the PaO2/FiO2 (PF) ratio, requirement for prone ventilation in the previous 24 hours or Murray lung injury score [17] between groups (p>0.36 for all, table 5).

#### Myocardial Injury

There was no difference in the *proportion* of patients with normal or abnormal NT-proBNP levels, between those with and without RVD (p=0.255). However, *median* NT-proBNP values were higher in those *with* RVD (p<0.006, table 5 and Figure 3). Conversely, there was no difference in *median* Troponin levels (I or T) between the groups (p>0.082). However, abnormal Troponin values were more frequent in those *with* RVD (p=0.048, table 5).

**Figure 3.**
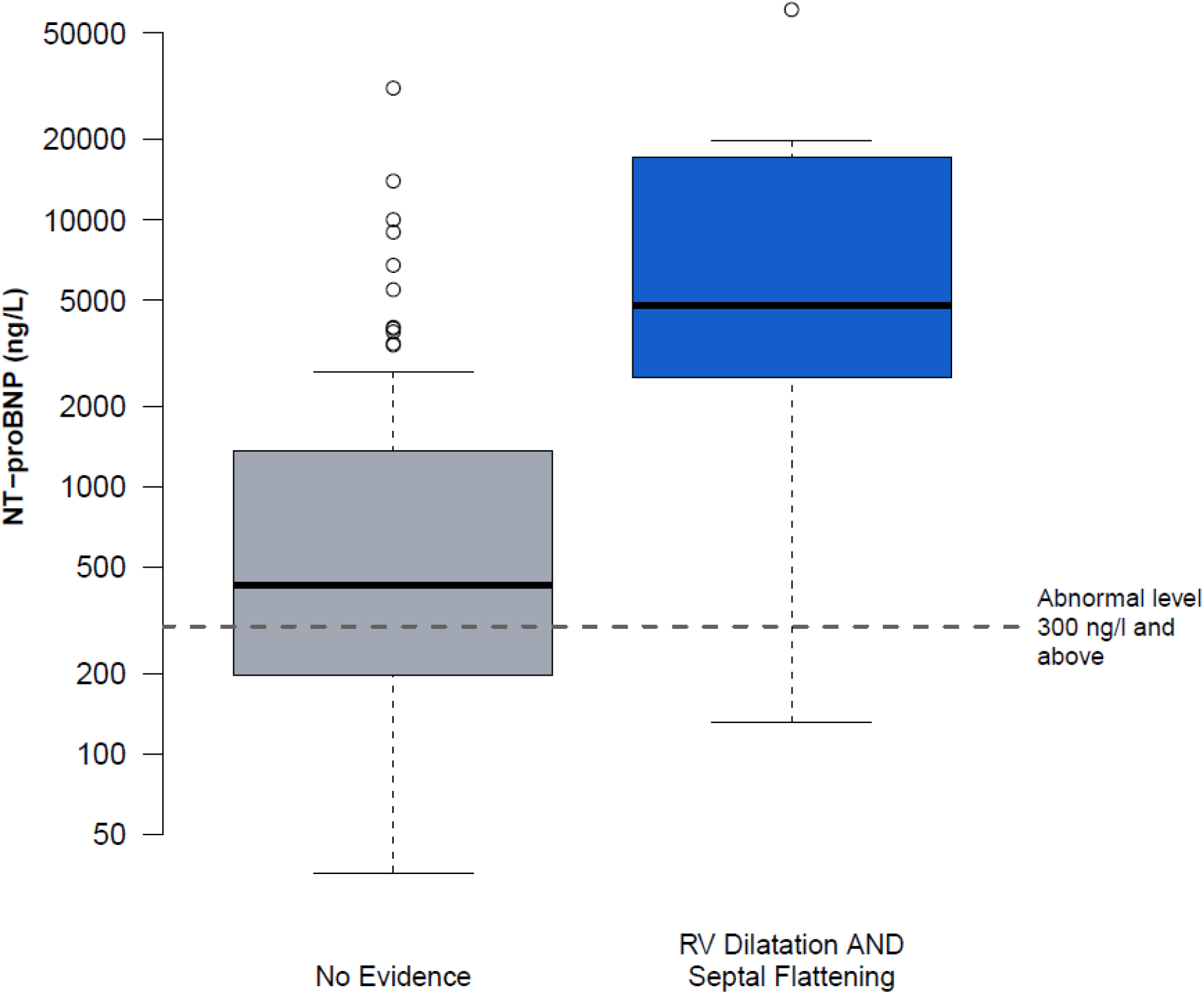
NT-proBNP levels in those with and without Right Ventricular Dysfunction (RV dilatation *and* septal flattening). NT-proBNP = N-terminal pro b-type natriuretic peptide.

Regression analyses controlling for demographics, phase of disease and baseline severity of illness demonstrated; an abnormal NT-proBNP (OR [95%CI] 4.77 [1.22, 21.32], p=0.029), and an abnormal Troponin (16.54 [4.98, 67.12], p<0.001) both independently predicted 30-day mortality (Table 6).

**Table 6.**
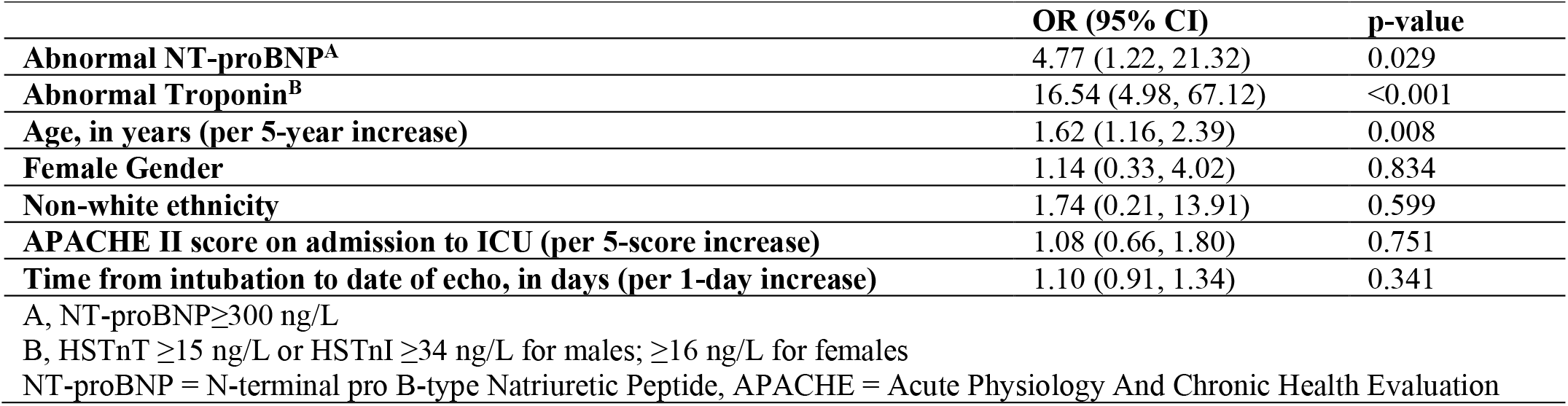
Logistic regression predicting 30-day mortality adjusting for remaining variables in table.

|  | **OR (95% CI)** | **p-value** |
| --- | --- | --- |
| **Abnormal NT-proBNP<sup>A</sup>** | 4.77 (1.22, 21.32) | 0.029 |
| **Abnormal Troponin<sup>B</sup>** | 16.54 (4.98, 67.12) | <0.001 |
| **Age, in years (per 5-year increase)** | 1.62 (1.16, 2.39) | 0.008 |
| **Female Gender** | 1.14 (0.33, 4.02) | 0.834 |
| **Non-white ethnicity** | 1.74 (0.21, 13.91) | 0.599 |
| **APACHE II score on admission to ICU (per 5-score increase)** | 1.08 (0.66, 1.80) | 0.751 |
| **Time from intubation to date of echo, in days (per 1-day increase)** | 1.10 (0.91, 1.34) | 0.341 |

A, NT-proBNP ≥ 300 ng/L
B, HSTnT ≥ 15 ng/L or HSTnI ≥ 34 ng/L for males; ≥ 16 ng/L for females
NT-proBNP = N-terminal pro B-type Natriuretic Peptide, APACHE = Acute Physiology And Chronic Health Evaluation

### Post-hoc analyses

Post-hoc analyses examining the alternative endpoint of RVD as defined by the presence of RV dilatation *and/or* septal flattening are presented as Supplementary tables. This endpoint occurred in 33 of 112 patients, 29.5% [95% CI; 21.2%, 38.8%], figure 1) but was not associated with mortality (51.5% in those with, compared to 45.6% in those without, p=0.679, supplementary table 4.). Regression analyses, controlling for patient demographics, phase of disease and baseline severity of illness demonstrated no association between this definition of RVD and 30-day mortality (Supplementary Table 6).

## DISCUSSION

To our knowledge, this is the first study to prospectively explore the prevalence of RV dysfunction in *ventilated patients* with COVID-19. The prevalence of RVD was 6.3% and this was associated with a mortality of 85.7%, in contrast to 44.8% in those without RVD.

A universal definition of *right ventricular dysfunction* is not well established; it has been suggested the term refers to structural changes (abnormal imaging and/or biomarkers) but with maintained cardiac output, a clinical setting which may progress to *right ventricular failure* (RVF). RVF is characterised by insufficient delivery of blood from the RV along with elevated systemic venous pressures [18]. As RVF is a difficult *clinical* diagnosis to make, we elected to refer to the echocardiographic changes as RVD. There is however, a high likelihood that patients in COVID-RV with a diagnosis of RVD would also have fitted clinical criteria for RVF, where there was associated evidence of tachycardia, systemic hypoperfusion (disordered acid-base status) and renal dysfunction.

Echocardiographic assessment of RV function is challenging; due to its complex geometry, retrosternal position, and marked load dependence. As a result, there is no gold-standard numeric measurement of RV function and international guidelines advocate assessment should incorporate a combination of both qualitative and quantitative parameters [19]. These quantitative methods however have been observed to vary markedly in their diagnostic performance [20]. More importantly, for COVID-RV, they are not assessed as part of a focused, bedside FICE echocardiogram. We deliberately selected a pragmatic definition of RVD, so as to empower bedside critical care clinicians, to make the diagnosis [21]. Identification of RVD could inform patient prognosis, alert to the potential for associated pathologies, such as PTE, and influence treatment strategies; such as alteration of ventilation, introduction of inotropic support or pulmonary vasodilators. Furthermore, should any interventional study of RV therapies be proposed in this patient group, it is imperative that any inclusion criteria (i.e., the diagnosis of RVD) allows the treating bedside clinician to make the diagnosis.

RV dilatation and septal flattening together constitute *acute cor pulmonale* (ACP). This has been more specifically defined as septal flattening with a dilated RV (RV:LV ratio >0.6, with >1.0 for *severe* ACP) [22]. The observed prevalence of RVD (defined as *severe* RV dilation *and* septal flattening) at 6.3% was lower than hypothesised. In the early phases of the pandemic, anecdotal clinical reports, social media posts and early peer reviewed literature suggested RVD was very common; RV dilatation was reported to be present in 31-39% of cases [1,3,4,23]. These studies however had mechanical ventilation rates of only 10-30% and therefore it was hypothesised the prevalence of RVD would be higher in a cohort requiring intubation and mechanical ventilation; previous reports of RVD in ARDS described a prevalence of up to 50% [13,24]. This lesser prevalence may have been observed for a number of reasons: Firstly, these early anecdotal, observational and retrospective reports are likely to have been influenced by selection bias, where only patients who were haemodynamically unstable or deteriorating would have imaging performed, meaning the prevalence of RVD was over reported. COVID-RV recruited 23.9% of all patients admitted to participating ICU’s during the study period; the prospective and systematic approach to patient recruitment, provides confidence that the prevalence of RVD presented is accurate. When compared to contemporaneous national reporting by both the Intensive Care National Audit & Research Centre (ICNARC) and the Scottish Intensive Care Society Audit Group (SICSAG), the patients recruited to COVID-RV are representative in terms of severity of illness, and mortality [25,26]. Secondly, there were significant differences in patient demographics, clinical care and outcomes between the time of these early reports and the UK’s “second wave” when the COVID-RV cohort was recruited. Attitudes to non-invasive respiratory support, ventilation practice, fluid management and anticoagulation all evolved and successful drug therapies were discovered and implemented; 66.1% of COVID-RV participants were treated with intravenous steroids prior to intubation [27,28]. Thirdly, the definition of RVD used in this study, necessitating the presence of both *severe* RV dilatation *and* septal flattening is a more stringent definition than used in other reports. The alternative endpoint of RV dilatation *and/or* septal flattening was evident in 29.5% of patients (Supplemental tables 3-6) whilst *subjective RV dysfunction* was diagnosed in 14.4%.

Our findings however, are in keeping with previous reports in ARDS. In a study of 752 patients by Mekontso Dessap et al. ACP was present in 22% (95% CI 19, 25%) and *severe* ACP (the same definition as used in COVID-RV) was present in 7.2% (95% CI 5.4%, 9.3%) [13]. Importantly, in this study, as in COVID-RV, hospital mortality was only higher in patients with *severe* ACP when compared with all other patients (57% compared to 42% in those without [p=0.03]), suggesting this definition of RVD identifies a population of patients where diagnosis yields significant clinical sequalae.

High driving pressure, low P/F ratio and high PaCO2 have been associated with RVD in patients with ARDS [13]. Although COVID-RV was not able reproduce these findings, the study demonstrated that plateau pressure was higher, and compliance lower, in those with RVD suggesting interplay between ARDS and the conduct of mechanical ventilation in the aetiology of RVD in this population.

The observed association between *confirmed or suspected* PTE and RVD is not un-expected and demonstrates the role of macrothrombi in RVD. It has been consistently demonstrated that patients with COVID-19 in ICU have a high frequency of thrombotic complications with a PTE incidence of 12.6% [29]. RVD is common in patients with PTE and is a major determinant of short-term survival [30,31].

Cardiomyocyte injury, quantified by elevated troponin levels, and haemodynamic cardiac stress, as quantified by increased natriuretic peptide concentrations have been previously described both in COVID-19 and ARDS [32–34]. The association of NT-proBNP and abnormal troponin levels with RVD in this cohort suggests a potential role of direct myocardial injury in the aetiology of myocardial dysfunction in patients with COVID-19. Our findings are consistent with previous work, with elevation of NT-proBNP and troponin strongly associated with mortality.

The lack of association between time from *intubation to echocardiography* and RVD suggests the phase of a patient’s *ICU* stay does not influence the presence of RVD. The timing of intubation however is multi-factorial, varying from patient to patient and ICU to ICU; some patients for example will be intubated late, following prolonged periods of non-invasive respiratory support, and others will have been intubated much earlier. However, the observed association between *time of symptom onset to echocardiography* and RVD, suggests the overall stage of a patient’s disease process may influence the presence of RVD. Previous work has demonstrated higher incidences of VTE with increasing time since COVID-19 diagnosis and given the association between PTE and RVD, this may contribute to this finding in the COVID-RV cohort [35].

Strengths of this study include a prospective, a-priori analysis of RVD in a cohort requiring mechanical ventilation using a well-established definition, known to be associated with outcome in patients with ARDS. A weakness of the study is the single timepoint of echocardiographic assessment, it is unknown whether patients classified as not having RVD may have had abnormal echocardiography if imaging had occurred at a different timepoint during their admission. Although an accurate report of the prevalence of RVD in ventilated patients with COVID-19, the low numbers of the primary outcome event prevent any in depth multi-variate assessment of the factors associated with RVD. In addition, the associations demonstrated are at risk of both type I and type II error, meaning they can only be considered exploratory in nature.

COVID-RV demonstrates that although the prevalence of RVD is lower than predicted in ventilated patients with COVID-19, it is associated with a high mortality. Association is observed between RVD and each of the aetiological domains of; ARDS, ventilation, micro/macrothrombi and myocardial injury. COVID-RV highlights the need for increased clinician awareness of RVD and should aid design of therapeutic studies seeking to improve outcome in this critically ill patient group.

## Data Availability

The data that support the findings of this study are available from the corresponding author, [BS], upon reasonable request and subject to appropriate ethical approvals.

