## Supplementary tables for "Right Ventricular Dysfunction in Ventilated Patients with COVID-19 (COVID-RV)"

| **Supplementary table 1. Indicative power** | | | | | | |
|---|---|---|---|---|---|---|
| **Sample Size** | **Power** | **Number *without* RVD** | **Number *with* RVD** | **Mortality in patients *without* RVD** | **Mortality in patients *with* RVD** | **Odds Ratio** |
| 120 | 80% | 60 | 60 | 0.373 | 0.627 | 2.83 |
| 120 | 90% | 60 | 60 | 0.354 | 0.646 | 3.33 |
| 150 | 80% | 75 | 75 | 0.386 | 0.614 | 2.53 |
| 150 | 90% | 75 | 75 | 0.369 | 0.631 | 2.92 |
| 120 | 80% | 90 | 30 | 0.427 | 0.719 | 3.43 |
| 120 | 90% | 90 | 30 | 0.417 | 0.749 | 4.17 |
| 150 | 80% | 112 | 38 | 0.435 | 0.695 | 2.96 |
| 150 | 90% | 112 | 38 | 0.426 | 0.722 | 3.50 |

Differences in mortality rates that could be detected at a 5% significance level for sample sizes of 120 and 150, and proportions of patients with RV dysfunction of 25% and 50%, assuming an overall mortality rate of 50%. RVD = Right Ventricular Dysfunction.

| Supplementary Table 2. Determination of primary outcome | | | | |
|---|---|---|---|---|
| **RV dilatation** | **Septal flattening** | **Primary outcome** | **Included/excluded** | **n** |
| Yes | Yes | Yes | Included | 7 |
| Yes | Unable to assess | Unknown | Excluded | 0 |
| Yes | No | No | Included | 24 |
| Unable to assess | No | No | Included | 2 |
| No | No | No | Included | 74 |
| No | Yes | No | Included | 2 |
| No | Unable to assess | No | Included | 3 |
| Unable to assess | Unable to assess | Unknown | Excluded | 6 |
| RV = Right Ventricle | | | | |

| Supplementary Table 3. Patient characteristics at ICU admission | | | | | | |
|---|---|---|---|---|---|---|
| | | | **All (n=112)** | **RVD (n=33)** | **No RVD (n=79)** | **p-value** |
| **Age, years** | | | 59.2 (11.3) | 60.7 (9.9) | 58.5 (11.8) | 0.326[#] |
| **Sex, Male** | | | 74 (66.1%) | 22 (66.7%) | 52 (65.8%) | >0.99[†] |
| **BMI, kg m-2** | | ***n (n missing)*** | *110 (2)* | 32 (1) | 78 (1) | |
| | | | 32.9 (7.1) | 32.9 (6.7) | 32.8 (7.3) | 0.952[#] |
| **Ethnicity** | **White** | | 100 (89.3%) | 26 (78.8%) | 74 (93.7%) | 0.039[†] |
| | **Non-white** | | 12 (10.7%) | 7 (21.2%) | 5 (6.3%) | |
| **Time since symptom onset to intubation, days** | | | 11 [7, 16] | 11.0 [7, 17] | 10.5 [7.2, 14] | 0.384[‡] |
| **Clinical frailty score** | | ***n (n missing)*** | *111 (1)* | 33 (0) | 78 (1) | |
| | | | 2.4 (0.9) | 2.4 (1) | 2.3 (0.9) | 0.930[#] |
| **APACHE II** | | ***n (n missing)*** | *107 (5)* | 33 (0) | 74 (5) | |
| | | | 16.6 (5.8) | 16.8 (5.1) | 16.6 (6.2) | 0.877[#] |
| **CCCC** | | ***n (n missing)*** | *102 (10)* | 29 (4) | 73 (6) | |
| | | | 10.2 (2.7) | 10.8 (1.9) | 10.0 (3) | 0.105[#] |
| ***Comorbidities*** | | | | | | |
| **Smoking** | **Non-smoker** | | 63 (56.2%) | 22 (66.7%) | 41 (51.9%) | |
| | **Ex-smoker > 1 year** | | 40 (35.7%) | 10 (30.3%) | 30 (38%) | 0.293[†] |
| | **Current or within 1 year** | | 9 (8%) | 1 (3%) | 8 (10.1%) | |
| **Alcohol history** | ***n (n missing)*** | | *110 (2)* | 33 (0) | 77 (2) | |
| | **None** | | 34 (30.9%) | 10 (30.3%) | 24 (31.2%) | |
| | **Minimal** | | 57 (51.8%) | 20 (60.6%) | 37 (48.1%) | 0.544[†] |
| | **Moderate** | | 8 (7.3%) | 1 (3%) | 7 (9.1%) | |
| | **Excess** | | 11 (10%) | 2 (6.1%) | 9 (11.7%) | |
| **Hypertension** | | | 38 (33.9%) | 13 (39.4%) | 25 (31.6%) | 0.513[†] |
| **Coronary artery disease** | | | 11 (9.8%) | 4 (12.1%) | 7 (8.9%) | 0.333[†] |
| **Diabetes** | | | 33 (29.5%) | 9 (27.3%) | 24 (30.4%) | 0.823[†] |
| **Asthma** | | | 16 (14.3%) | 4 (12.1%) | 12 (15.2%) | 0.775[†] |
| **COPD** | | | 10 (8.9%) | 2 (6.1%) | 8 (10.1%) | 0.803[†] |
| ***Treatments before intubation*** | | | | | | |
| **Intravenous corticosteroids** | | | 74 (66.1%) | 25 (75.8%) | 49 (62%) | 0.193[†] |
| **Non-invasive ventilation** | | | 76 (67.9%) | 22 (66.7%) | 54 (68.4%) | >0.99[†] |
| **High flow nasal oxygen** | | | 65 (58%) | 24 (72.7%) | 41 (51.9%) | 0.058[†] |
| **Awake self-proning** | | | 57 (50.9%) | 19 (57.6%) | 38 (48.1%) | 0.411[†] |
| ***Acute comorbidities since hospital admission*** | | | | | | |
| **New arrhythmias** | | | 17 (15.2%) | 6 (18.2%) | 11 (13.9%) | 0.573[†] |
| **Confirmed or suspected PTE** | **Radiologically confirmed** | | 4 (3.6%) | 4 (12.1%) | 0 (0%) | |
| | **Clinically suspected** | | 5 (4.5%) | 2 (6.1%) | 3 (3.8%) | 0.010[†] |
| | **No** | | 101 (90.2%) | 27 (81.8%) | 74 (93.7%) | |
| | **Unknown** | | 2 (1.8%) | 0 (0%) | 2 (2.5%) | |
| **ACS** | | | 5 (4.5%) | 2 (6.1%) | 3 (3.8%) | 0.630[†] |
| **Requirement for RRT** | | | 18 (16.1%) | 6 (18.2%) | 12 (15.2%) | 0.779[†] |

RVD defined as RV dilatation *and/or* septal flattening

Data are presented as mean (SD), median [IQR] or n (%). Data are complete unless indicated by *n (n missing)*. P-values provided are for comparisons between RVD vs no RVD. # = Student's T-test. † = Fisher's Exact test. ‡ = Wilcoxon Mann-Whitney test.

RVD = Right Ventricular Dysfunction, BMI = Body Mass Index, APACHE = Acute Physiology And Chronic Health Evaluation, CCCC = Coronavirus Clinical Characterisation Consortium, COPD = Chronic Obstructive Pulmonary Disease, PTE = Pulmonary Thromboembolism, ACS = Acute Coronary Syndrome, RRT = Renal Replacement Therapy.

**Supplementary Table 4. Clinical consequences**

| | All (n=112) | RVD (n=33) | No RVD (n=79) | p-value |
|---|---|---|---|---|
| ***30-day follow-up*** | | | | |
| **Death** | 53 (47.3%) | 17 (51.5%) | 36 (45.6%) | 0.679† |
| **RRT** | 28 (25%) | 9 (27.3%) | 19 (24.1%) | 0.879† |
| **Prone ventilation** | 55 (49.1%) | 13 (39.4%) | 42 (53.2%) | 0.213† |
| **Referral for ECMO** | 15 (13.4%) | 1 (3.0%) | 14 (17.7%) | 0.097† |

RVD defined as RV dilatation *and/or* septal flattening

Data are presented as n (%). Data are complete unless indicated by *n (n missing).* P-values provided are for comparisons between RVD vs no RVD. † = Fisher's Exact test.

RRT = Renal Replacement Therapy, ECMO = Extra Corporeal Membrane Oxygenation.

**Supplementary Table 5. Patient characteristics on day of echocardiography**

| | | All (n=112) | RVD (n=33) | No RVD (n=79) | p-value |
|---|---|---|---|---|---|
| **Requirement for prone invasive ventilation since ICU admission** | | 79 (70.5%) | 28 (84.8%) | 51 (64.6%) | 0.061† |
| **SOFA score** | | 7.9 (3) | 8.8 (3.2) | 7.6 (2.8) | 0.048# |
| **Requirement for RRT on day of echo** | | 15 (13.4%) | 7 (21.2%) | 8 (10.1%) | 0.135† |
| ***Lab Measurements*** | | | | | |
| ***Arterial Blood Gas*** | | | | | |
| **$[H^+]$, nmol $L^{-1}$** | ***n (n missing)*** | *97 (15)* | 29 (4) | 68 (11) | |
| | | 39 [35.8, 46] | 40.9 [36, 49] | 39 [34.8, 45] | 0.230‡ |
| **$PaO_2$, kPa** | | 9.3 (1.2) | 9.1 (1.3) | 9.4 (1.2) | 0.428# |
| **$PaCO_2$, kPa** | ***n (n missing)*** | *109 (3)* | 33 (0) | 76 (3) | |
| | | 6.9 [5.9, 8] | 7.0 [5.9, 7.9] | 6.8 [5.9, 8] | 0.929‡ |
| **BE, mmol** | ***n (n missing)*** | *110 (2)* | 33 (0) | 77 (2) | |
| | | 5.9 (6.5) | 3.8 (6.4) | 6.8 (6.3) | 0.026# |
| **Bicarbonate, mmol $L^{-1}$** | ***n (n missing)*** | *109 (3)* | 33 (0) | 76 (3) | |
| | | 31.8 (6.6) | 30.0 (6.1) | 32.5 (6.7) | 0.063# |
| ***Full Blood Count*** | | | | | |
| **Haemoglobin, g $dl^{-1}$** | | 11 (1.8) | 10.7 (1.4) | 11.2 (1.9) | 0.127# |
| **Neutrophils, $x10^9$ $L^{-1}$** | | 10.5 [8.5, 14.9] | 12.3 [8.9, 17.7] | 10.3 [8.4, 13.6] | 0.128‡ |
| **Lymphocytes, $x10^9$ $L^{-1}$** | | 0.9 [0.5, 1.4] | 0.9 [0.7, 1.5] | 0.9 [0.4, 1.3] | 0.239‡ |
| **Platelets, $x10^9$ $L^{-1}$** | | 279.5 (109.5) | 276.5 (105.7) | 280.8 (111.7) | 0.849# |
| ***Inflammation*** | | | | | |
| **CRP, mg $L^{-1}$** | | 61.5 [11.8, 157.5] | 72.0 [14, 178] | 54.0 [11, 149] | 0.375‡ |
| ***Coagulation*** | | | | | |
| **D-Dimers, mg $L^{-1}$ FEU** | ***n (n missing)*** | *81 (31)* | 27 (6) | 54 (25) | |
| | | 1264 [601, 2605] | 1156 [504, 2822] | 1373 [724.8, 2496] | 0.745‡ |
| **PT, seconds** | | 12 [11, 13.2] | 13 [11, 13] | 12 [11, 13.2] | 0.264‡ |
| **APTT, seconds** | ***n (n missing)*** | *111 (1)* | 33 (0) | 78 (1) | |
| | | 27 [25, 31] | 29 [26, 31] | 26 [24, 30] | 0.031‡ |
| ***Electrolytes*** | | | | | |
| **Creatinine, µmol $L^{-1}$** | | 69.5 [53.5, 107] | 84 [54, 118] | 64 [53, 99] | 0.099‡ |
| ***Cardiac biomarkers*** | | | | | |
| **NT-proBNP, ng $L^{-1}$** | ***n (n missing)*** | *100 (12)]* | 32 (1) | 68 (11) | |
| | | 458 [198.5, 1689.5] | 537.5 [208, 3634.2] | 430.5 [194, 1275.2] | 0.213‡ |
| *Abnormal* **NT-proBNP[A]** | ***n (n missing)*** | *100 (12)* | 32 (1) | 68 (11) | |
| | | 63 (63%) | 20 (62.5%) | 43 (63.2%) | >0.99† |
| **hsTn I, ng $L^{-1}$** | ***n (n missing)*** | *64 (48)* | 24 (9) | 40 (39) | |
| | | 12[4.8, 42] | 19.5 [4.8, 42] | 10.0 [4.8, 40] | 0.605‡ |
| **hsTn T, ng $L^{-1}$** | ***n (n missing)*** | *46 (66)* | 9 (24) | 37 (42) | |
| | | 16.5 [10, 28.5] | 18 [15, 37] | 16 [10, 26] | 0.239‡ |
| *Abnormal* **troponin[B]** | ***n (n missing)*** | *110 (2)* | 33 (0) | 77 (2) | |
| | | 51 (46.4%) | 17 (51.5%) | 34 (44.2%) | 0.535† |
| ***Clinical Parameters*** | | | | | |
| **HR, bpm** | | 79 (19.9) | 81.3 (21.1) | 78.1 (19.5) | 0.446# |
| **Rhythm** | **Sinus** | 107 (95.5%) | 31 (93.9%) | 76 (96.2%) | |
| | **AF/Flutter** | 5 (4.5%) | 2 (6.1%) | 3 (3.8%) | 0.630† |
| **Mean BP, mmHg** | ***n (n missing)*** | *109 (3)* | 32 (1) | 77 (2) | |
| | | 80.9 (13.4) | 77.9 (10.5) | 82.1 (14.3) | 0.089# |
| **CVP, $cmH_2O$** | ***n (n missing)*** | *74 (38)* | 22 (11) | 52 (27) | 0.307‡ |
| | | 7 [4, 12] | 7.5 [6.0, 13.8] | 7.0 [3.8, 11.2] | |
| ***Drug Administration*** | | | | | |
| **Vasopressors** | | 40 (35.7%) | 18 (54.5%) | 22 (27.8%) | 0.010† |
| **Inotropes** | | 1 (0.9%) | 1 (3%) | 0 (0%) | 0.295† |
| **Anticoagulation** | **Prophylactic** | 93 (83%) | 24 (72.7%) | 69 (87.3%) | |
| | **Therapeutic** | 17 (15.2%) | 9 (27.3%) | 8 (10.1%) | 0.057† |
| | **None** | 2 (1.8%) | 0 (0%) | 0 (0%) | |
| **Paralysis** | | 52 (46.4%) | 18 (54.5%) | 34 (43%) | 0.511† |
| ***Ventilation*** | | | | | |
| **$FiO_2$** | | 0.6 [0.4, 0.7] | 0.6 [0.5, 0.7] | 0.6 [0.4, 0.7] | 0.083‡ |
| **Requirement for prone ventilation in previous 24 hours** | | 44 (39.3%) | 16 (48.5%) | 28 (35.4%) | 0.498† |

| | | | | | |
|---|---|---|---|---|---|
| **Plateau pressure, $cmH_2O$** | | 58 (54) | 19 (14) | 39 (40) | |
| | | 24 [22, 27] | 27 [25, 29.5] | 24 [22, 26] | 0.018[‡] |
| **PAP, $cmH_2O$** | ***n (n missing)*** | *58 (54)* | 33 (0) | 76 (3) | |
| | | 25 [20, 30] | 27 [21, 30] | 24 [20, 28.2] | 0.176[‡] |
| **Tidal volume, ml $kg^{-1}$** | ***n (n missing)*** | *108 (4)* | 32 (1) | 76 (3) | |
| **(PBW)** | | 7.2 (2.1) | 7.2 (2.5) | 7.2 (2) | 0.964[#] |
| **P/F ratio** | | 17 [13.6, 21.3] | 15.2 [11.7, 18] | 17.8 [14.3, 21.4] | 0.047[‡] |
| **PEEP, $cmH_2O$** | ***n (n missing)*** | *111 (1)* | 33 (0) | 78 (1) | |
| | | 9.8 (3.6) | 10.2 (3.4) | 9.7 (3.7) | 0.447[#] |
| **Resp rate, $minute^{-1}$** | | 24.3 (5.3) | 24.8 (5.2) | 24.1 (5.3) | 0.489[#] |
| **Driving pressure, $cmH_2O$** | ***n (n missing)*** | *58 (54)* | 19 (14) | 39 (40) | |
| | | 14.7 (7.1) | 15.9 (5.7) | 14.0 (7.6) | 0.290[#] |
| **Dynamic compliance, ml** | ***n (n missing)*** | *58 (54)* | 19 (14) | 39 (40) | |
| **$cmH_2O^{-1}$** | | 29.8 [20, 40] | 21.3 [17.9, 36] | 34.3 [22.7, 41.8] | 0.082[‡] |
| **Murray lung injury score** | ***n (n missing)*** | 100 (12) | 31 (2) | 69 (10) | |
| | | 2.8 [2.3, 3] | 2.8 [2.5, 3.3] | 2.8 [2.3, 3] | 0.491[‡] |

RVD defined as RV dilatation *and/or* septal flattening

Data are presented as mean (SD), median [IQR] or n (%). Data are complete unless indicated by *n (n missing).* P-values provided are for comparisons between RVD vs no RVD. # = Student's T-test. † = Fisher's Exact test. ‡ = Wilcoxon Mann-Whitney test. Missing data indicated by *n (n missing).*

A, NT-proBNP ≥300 ng $L^{-1}$

B, hsTnT ≥15 ng $L^{-1}$ or hsTnI ≥34 ng $L^{-1}$ for males; ≥16 ng $L^{-1}$ for females

RVD = Right Ventricular Dysfunction, RRT = Renal Replacement Therapy, BE = Base Excess, CRP = C-Reactive Protein, PT = Prothrombin Time, APTT = Activated Partial Thromboplastin Time, NT-proBNP = N-terminal pro B-type Natriuretic Peptide, hsTn = High Sensitivity Troponin, HR = Heart Rate, AF = Atrial Fibrillation, BP = Blood Pressure, CVP = Central Venous Pressure, $FiO_2$ = Fraction of Inspired Oxygen, PAP = Peak Airway Pressure, PBW = Predicted Body Weight, PEEP = Positive End Expiratory Pressure.

| **Supplementary Table 6. Logistic regression model predicting 30-day mortality adjusting for remaining variables in table** | | |
|---|---|---|
| | **OR (95% CI)** | **p-value** |
| **RV dilatation *and/or* septal flattening** | 1.35 (0.54, 3.46), | =0.523 |
| **Age, in years (per 5-year increase)** | 1.52 (1.21, 1.97), | =0.001 |
| **Female Gender** | 1.09 (0.44, 2.70), | =0.855 |
| **Non-white ethnicity** | 0.68 (0.15, 2.90), | =0.610 |
| **APACHE II score on admission to ICU (per 5-score increase)** | 1.30 (0.88, 1.98), | =0.191 |
| **Time from intubation to date of echo, in days (per 1-day increase)** | 1.02 (0.88, 1.17), | =0.826 |
| APACHE = Acute Physiology And Chronic Health Evaluation | | |
